# Association of DOT1L histone methyltransferase with non-syndromic orofacial clefts in a cohort of children of African and Asian ancestry

**DOI:** 10.64898/2026.06.26.26356682

**Authors:** Timothy Becker, Dashzeveg Bayarsaihan, Dong-Guk Shin

## Abstract

Non-syndromic orofacial clefts (nsOFCs), such as cleft lip, cleft lip with or without cleft palate (CL/P), and cleft palate only, are among the most common craniofacial malformations in humans. We identified a range of rare and *de novo* structural variants (SVs) associated with nsOFCs in a cohort of children of African and Asian ancestry from the Gabriella Miller Kids First Data Resource Center. Most of these novel candidate SVs are located in non-coding regions. More specifically, we characterized SVs associated with nsOFCs across the *DOT1L* (disruptor of telomeric silencing 1-like) genomic locus. DOT1L is the sole histone methyltransferase that catalyzes the mono-, di-, and trimethylation of histone H3 at lysine 79 (H3K79). In addition, our data revealed associations between specific SVs and genes encoding DOT1L complex subunits as well as downstream target genes. Single-nuclear RNA sequencing analysis demonstrated that mouse *Dot1L* and its associated targets are highly expressed in neural crest-derived mesenchyme enriched for key osteogenic genes essential for palate formation. Depletion of a *Dot1L* allele resulted in cleft palate and micrognathia in mice. Furthermore, we identified several families carrying SVs in the *DOT1L* locus and downstream DOT1L targets genes. These findings support an oligogenic model in which the concurrent presence of several SVs enhances susceptibility to the nsOFC phenotype. We propose that DOT1L and its targets act within the same genetic pathways to influence CL/P such that the combined effects of variants affecting these genes may be more substantial than their individual effects. Collectively, these studies establish a pivotal role for DOT1L-mediated H3K79 methylation in craniofacial development and palate formation and identify this pathway as a key contributor to the etiology of CL/P.

## Introduction

Non-syndromic orofacial clefts (nsOFCs), including cleft lip, cleft lip with or without cleft palate (CL/P), and cleft palate only, are among the most common craniofacial malformations in humans (Candotto et al., 2019). nsOFCs affect approximately 1 in 700 live births worldwide, with significant variation among populations (Babai et al., 2023). Although emerging evidence highlights the crucial role of epigenetic regulation in the occurrence and progression of CL/P (Im et al., 2025; Alade et al., 2022; Shull et al., 2024), the mechanisms by which specific epigenetic modifications regulate distinct gene networks remain poorly understood. Epigenetic alterations in chromatin can directly influence transcriptional output, revealing the interplay between *cis*-regulatory DNA elements and target gene expression. It is well established that environmental risk factors, including maternal smoking, alcohol exposure, retinoic acid, and folate deficiency interact with epigenetic mechanisms and contribute to the complex etiology of nsOFCs (Im et al., 2025). The highly conserved histone methyltransferase disruptor of telomeric silencing 1-like (DOT1L) is enriched at actively transcribed genes through its interaction with the phosphorylated C-terminal domain of RNA polymerase II (Steger et al., 2008; Kim et al., 2012). DOT1L is the sole enzyme that catalyzes the mono-, di-, and trimethylation of histone H3 at lysine 79 (H3K79) (Feng et al., 2002). The enrichment of H3K79me2/3 within gene bodies correlates positively with active transcription (Wille et al., 2023). Notably, genome-wide analyses have shown that H3K79me2 is deposited at *cis*-regulatory elements, including enhancers and promoters (Wood et al., 2018; Ferrari et al., 2020). However, not all actively expressed genes are marked by H3K79me2, as DOT1L associates with lineage-specific genes in a context-dependent manner (Nguyen and Zhang, 2011). Another study suggests that DOT1L may also regulate transcriptional initiation by recruiting transcription factor IID (Wu et al., 2021). During neuronal differentiation of murine embryonic stem cells, accumulation of H3K79me2 is associated with SOX2-bound enhancers (Ferrari et al., 2020). Moreover, H3K79me2 is essential for maintaining enhancer–promoter interactions in leukemia cells (Godfrey et al., 2019).

The DOT1L complex includes co-factors such as SIRT1, MENIN, MLLT3, MLLT6, MLLT10, BCOR, and the WNT signaling components TRRAP, SKP1, and β-catenin (Mohan et al., 2010). This complex is thought to facilitate gene transcription by directly recruiting RNA polymerase II or by silencing repressive complexes containing HDACs and SUV39H1 (Guzzo et al., 2022). The essential role of DOT1L in mammalian development is well documented. Germline disruption of *Dot1L* and loss of H3K79me2 in mice lead to multiple developmental abnormalities, including growth impairment, angiogenesis defects in the yolk sac, cardiac dilation, and mid-gestational lethality due to defective extraembryonic vascular formation and impaired hematopoietic development (Jones et al., 2008; Feng et al., 2010). We previously demonstrated that mesenchyme-specific loss of *Dot1L* results in a skeletal dysplasia phenotype in mice (Sutter et al., 2021). Our research further implicated DOT1L in regulating key signaling pathways and core genes involved in proliferation and cell cycle checkpoint control in growth plate chondrocytes.

Notably, several DOT1L co-factors also play critical roles in craniofacial development. Deletion of *Menin* in cranial neural crest cells (cNCCs) causes cranial bone defects, including cleft palate (Engleka et al., 2007). Inactivation of *Bcor* in cNCCs results in cleft palate and shortening of the mandible and tympanic bone (Hamline et al., 2020). Deletion of *Mllt10* leads to midline facial cleft and ocular hypertelorism (Ogoh et al., 2017). Importantly, *TFAP2A*, a gene crucial for midfacial development, is directly regulated by MLLT10-dependent H3K79 methylation (Ogoh et al., 2022). Collectively, these studies strongly support a pivotal role for DOT1L-mediated H3K79 methylation in orofacial development and palate formation. However, whether structural variants (SVs) affecting DOT1L and its regulatory network contribute to nsOFC susceptibility remains largely unknown.

In this study, we investigated rare and *de novo* OFC-associated SVs in the *DOT1L* locus in a cohort of children of African and Asian ancestry from the Gabriella Miller Kids First Data Resource Center. Because many of the identified SVs localized in the regulatory regions of *DOT1L*, we further analyzed SVs associated with DOT1L co-factors and downstream target genes to define the contribution of the DOT1L regulatory network to nsOFCs.

## Materials and Methods

### Bioinformatics and computational approaches

#### Rare and de novo SVs (minor allele frequency, − MAF)

All SVs were clustered by type (DEL, DUP, INV, INS) using the FusorSV framework after integration with a 0.75 reciprocal overlap threshold. These clusters were then used to calculate allele frequencies in the background dataset (1000 Genome Project Phase 4; G1KP4). Allele frequencies below 5% in the background population were defined as rare, whereas SVs absent from the background dataset were assigned a frequency of 0% and considered *de novo*. In addition, we counted the number of samples from the OFC cohort (parents and children) to determine allele frequencies separately within the parental and affected-child groups.

#### SV segmental function score (SV-function)

We computed segmental functional scores using deleterious SVs (deletions and insertions) identified in the G1KP4 population by integrating the boundaries of all FusorSV per-sample breakpoint calls and assigning segmental functional score (SV-function) as follows: 1.0 − (count/total). A score of 1.0 indicates a highly-conserved genomic region lacking SV segments in the background population, whereas a score of 0.0 indicates that all background samples contain an SV segment at that location. By aggregating these scores and determining the frequency of each segment for each SV type, we generated a probability distribution function for SV segments across the reference coordinate space. This calculation provides a more accurate measure of individual genomic locations than allele frequency, which traditionally uses SV events where the reciprocal overlap exceeds 50%. Because our method integrates information at the base-pair level, it does not rely on reciprocal overlap for this distribution. This allows for greater accuracy in calculations that would involve 25% reciprocal overlap for example.

#### Cell-specific score (Cell-score)

We downloaded ChIP-seq peak files for H3K4me1 (poised enhancer mark), H3K4me3 (promoter mark), H3K27ac (active enhancer mark), and CTCF (insulator mark) from ENCODE for all available cell types. This comprised the embryonic stem cell line H9 and differentiated hepatocytes, mesenchymal stem cells (MSCs), mesothelial epicardial cells, nephron progenitors, NCCs, neural progenitors, osteocytes, pancreatic cells, peripheral blood cells, smooth muscle cells, spleen cells, and stomach cells. Only these 13 cell types had enhancer, promoter and insulator data available, whereas the remaining 12 of the total 25 available cell types either lacked histone peak data or were neural specific (e.g., brain). Together, the selected cells were representatives of the three major developmental branches (endoderm, mesoderm, and ectoderm) and were used to build a cell-specific score (Cell-score) through integration of individual peak breakpoint segments according to the formula 1− (count/total). Under this framework, a peak segment unique to a single cell type receives a score 0.92, whereas a segment shared across all cell types receives a score of 0.0. This serves as an estimate of the cell-specific probability distribution to determine where cell types are uniquely active relative to all other cells.

#### Transcription factor binding sites (TFBS) and promoter candidates

We used the JASPAR database, which contains a large collection of position weight matrices that enable TFBS prediction on sequence reads or RefSeq loci to determine the binding potential of any DNA sequence. We used the pyjaspar Python package (https://github.com/asntech/pyjaspar) to compute a normalized binding score for each transcription factor.

We computed distances from the transcription start site (TSS) using the first exon of the RefSeq Known Gene annotation table for hg38 (https://hgdownload.cse.ucsc.edu/goldenpath/hg38/bigZips/genes/) and H3K27me3 histone peaks. We used a 3 kb upstream search to identify H3k27me3 peaks within the range of each TSS and establish promoter candidate regions for each cell type.

#### Enhancer candidates and mutual proximity score (AEC, PEC, and MP-score)

We used H3K27ac peaks to identify promoter and enhancer candidates within the same cell type across a 1 Mbp search − window. We linked each active and poised enhancer peak to each promoter peak candidate and calculated the mutual distance ranking for each potential pairing. The harmonic mean of the mutual distance ranks was used to determine the enhancer-promoter mutual proximity score (MP-score), where 1.0 indicates the closest enhancer-promoter pairing and 0.0 indicates the most distant pairing among those possible within the 1 Mbp search window. We used ENCODE Hi-C preprocessed BED data (https://github.com/ENCODE-DCC/hic-pipeline) for several cell types, including NCC, MSCs, cerebellar astrocytes, motor neurons, B cells, pancreatic cells, heart ventricle cells, liver cells, kidney cells, spleen cells, and stomach cells.

#### Association of SVs with genes (direct SV-gene scoring)

In general, SVs received an association score of 0.0 for each gene. We then created associations using established functional rules based on the in-gene scoring (Danis et al., 2022). For direct SV-gene scoring, we used the same mechanism as SVAnna. For in-gene SVs, deletions or insertions affecting one or more exons received a pathogenetic score of 1.0. Duplications that encompass only some exons but not the entire gene (partial gene duplications) received a score of 1.0. Likewise, inversions affecting only a subset of exons received a score of 1.0. In SVAnna, introns were scored as 0.0, which is where our extended regulatory scoring framework diverges.

#### Association with genes (regulatory SV-gene scoring)

Because we have integrated histone marks, TFBSs, and Hi-C loop information, we also created indirect associations with SVs that lie outside the traditional gene region as defined by Refseq (hg38 in our study). In particular, we used the complete pairing information described above, termed the mutual proximity score, to score SVs that intersect promoter regions as: number of affected promoter candidates / total promoter candidates associated with a gene (within the 3 kb upstream search radius). Poised enhancers were scored as: number of affected poised enhancer candidates / total poised enhancer candidates within range of the promoter region. The active enhancers are scored as: number of active enhancer candidates / total poised enhancer candidates in range of the promoter region. Enhancers also carry an MP-score, which provide additional context regarding the proximity of these associations. Only deletion and insertion SVs received non-zero scores because it is not well established whether, and to what extent, an inverted or duplicated enhancer may affect transcript regulation. In these cases, we used a small non-zero value to indicate that the SV has some association with the regulatory element. Because regulatory marks commonly occur within intronic regions, our approach enables scoring across both gene bodies and associated regulatory regions.

### Animal work

We conditionally inactivated *Dot1L* in cNCCs using *Dot1L^fl/fl^* and *Wnt1-Cre2* mice from JAX Labs (129S4.Cg-^E2f1Tg(Wnt1-cre)2Sor^/J, strain #:022137). *Dot1L^fl/fl^* mice carry loxP sites flanking exon 5, which encodes most of the H3K79 methyltransferase domain (Sutter et al., 2021). We crossed female *Dot1L^fl/fl^* mice with *Wnt1*-*Cre2* mice, and the resulting *Dot1L^fl/+^;Wnt1-Cre* male offspring with *Dot1L^fl/fl^* females to produce conditional knockout *Dot1L^fl/fl^;Wnt1-Cre* (*Dot1L* cKO) mice. We performed PCR analysis according to the previously published protocol (Sutter et al., 2021). Skeletal preparations of mouse embryos at embryonic day 17.5 (E17.5) were performed according to previously published protocols (Baffi et al., 2004).

### Single-nucleus RNA sequencing (snRNA-seq)

#### 3’ Single Cell Library Preparation and Sequencing

Nuclei were isolated from flash-frozen E12.5 mouse embryonic heads according to the 10xGenomics cell preparation guide (Document CG00053, 10xGenomics.com). After dissociation, nuclei were washed and suspended in PBS containing 2% BSA and immediately processed. Nuclei viability was assessed on a LUNA FX7 automated cell counter (Logos Biosystems), and up to 14,420 nuclei from each suspension were loaded onto one lane of a 10x Genomics GEM-X 3’ Chip. Single cell capture, barcoding and library preparation were performed using the 10x Genomics Chromium X platform (https://www.nature.com/articles/ncomms14049) version 4.0 GEM-X chemistry, according to the manufacturer’s protocol (#CG000731 Rev B). cDNA and libraries were checked for quality using a TapeStation 4200 (Agilent) and Qubit Fluorometer (Thermo Fisher), quantified by KAPA qPCR, and sequenced on an Illumina NovaSeq X+ 10B 100- or 200-cycle flow cell lane, with a 28-10-10-90 asymmetric read configuration, targeting 10,000 barcoded nuclei at an average sequencing depth of 50,000 reads per nucleus. Illumina base call files for all libraries were converted to FASTQs using bcl2fastq v2.20.0.422 (Illumina), and FASTQ files associated with the gene expression libraries were aligned to the GRCm39 reference genome with GENCODE VM33 annotations (10x Genomics reference package 2024-A) using the Cell Ranger Multi pipeline version 9.0.1 (10x Genomics).

## Results

### The presence of rare and *de novo* SVs in the *DOT1L* region

Genomic variation encompasses both single-nucleotide variants (SNVs) and large changes (50 bp or greater) known as SVs. While the Kids First Data Resource Center has already completed SNV analysis, the goal of our research was to perform the complementary SV analysis needed to complete the comprehensive genomic landscape of the whole−genome sequencing (WGS) datasets. SVs are significantly larger than SNVs and, therefore exert substantially greater effects on gene networks, potentially advancing our understanding of the etiology of nsOFCs. We used 30× mean coverage Illumina paired-end WGS data from Kids First and the G1KP4 dataset to perform SV analysis. These data are suitable for gold −standard SNV analysis and allow detection of a wide variety of genomic aberrations. Using this holistic approach, we characterized the variability and ranked all of the DOT1L complex subunit regions (Mohan et al., 2010) with respect to conserved and variation-prone regions in healthy human populations and nsOFC populations. We analyzed 241 parent–child trios from the African and Asian Orofacial Clefts Case–Parent Triads datasets, along with a larger cohort of 3,000 healthy genomes representing 26 populations. In addition, we integrated SVs within DOT1L complex subunit regions with SVs in DOT1L downstream target genes (Sutter et al., 2021).

Our SV analysis followed a comprehensive and robust protocol comprising eight specialized SV callers that power an ensemble framework called FusorSV (Becker et al., 2018). Using this system, we can detect a wide range of SVs from 50 bp to more than 100 Mbp, including whole-chromosomal aneuploidies. This includes deletions, insertions, duplications, and inversions, listed in order of highest to lowest occurrence in human populations. In general, the ensemble uses at least two callers for each subtype; MELT is designed for insertion detection, whereas CNVnator is used for large deletion and duplication calling. FusorSV uses previously validated SVs (supervision) to learn the optimal combinations and cutoffs for removing overlapping or low-confidence calls, producing merged results with greater than 80% *in vivo* validation accuracy (Becker et al., 2018). We defined “rare SVs” as those occurring in less than 1% of the G1KP4 population and “*de novo* SVs” as rare variants that were absent from unaffected parent group. Collectively, we assembled all rare and *de novo* SVs in DOT1L complex subunits and DOT1L target genes associated with the nsOFC phenotype.

Within the genomic region encompassing *DOT1L*, we identified eight *de novo* SVs in eight CL/P probands that were absent in the unaffected parents (Table 1). Some of these SVs reside in the regulatory regions such as enhancers and promoters. In addition, we identified five *de novo* SVs associated with four DOT1L subunits, *ALYFREF, CSNK2A1*, *MLLT3* and *MLLT6*, in the probands (Table 2).

**Table 1.**
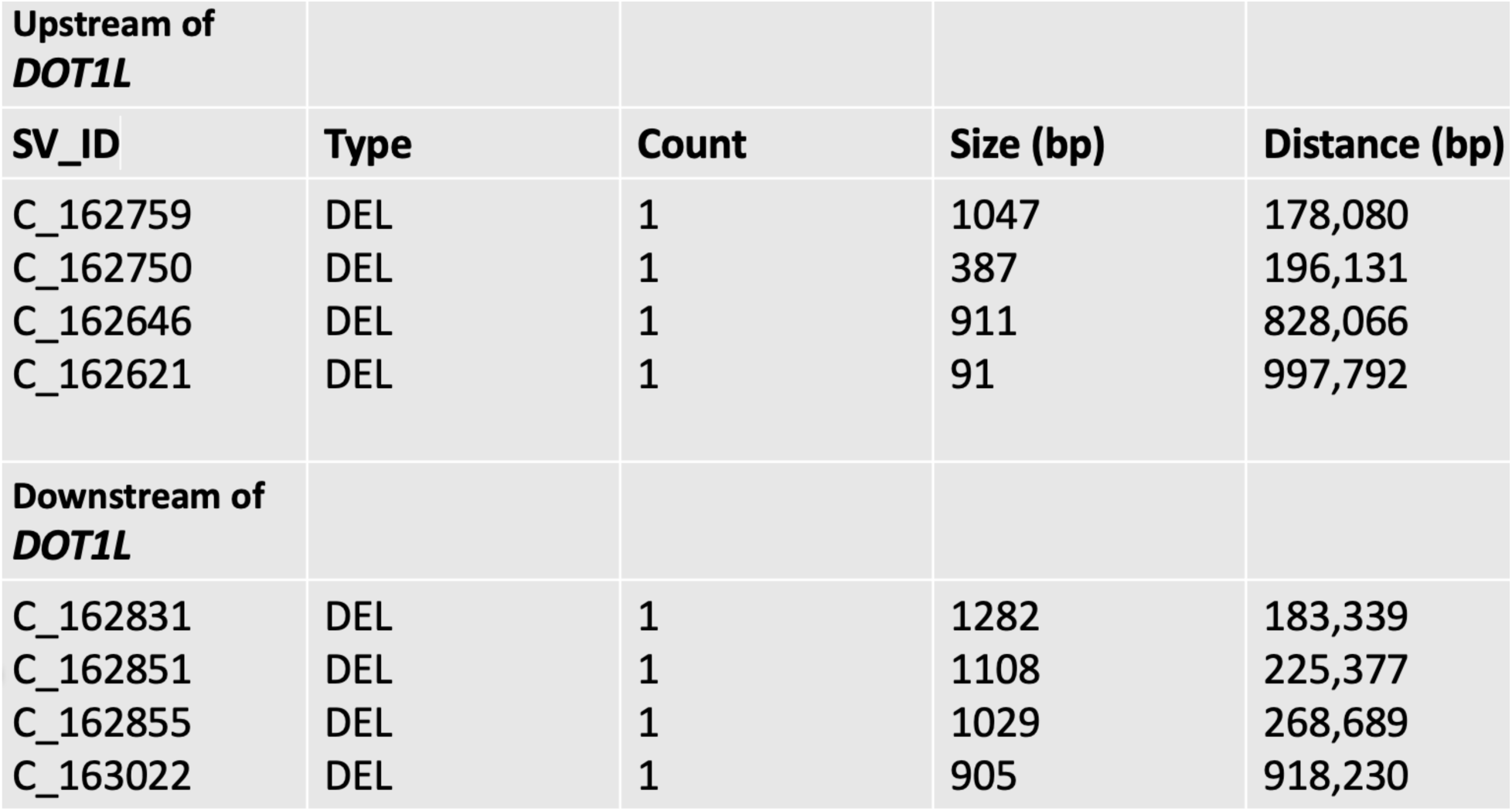
In the *DOT1L* region, we identified eight *de novo* SVs in eight nsOFC child samples that were absent in parents with normal phenotypes. DEL, deletion; count, number of affected children; distance, genomic distance in bp from the TSS of *DOT1L*; status, *de novo* SVs.

|  |  |  |  |  |
| --- | --- | --- | --- | --- |
| **Upstream of**<br>***DOT1L*** |  |  |  |  |
| **SV_ID** | **Type** | **Count** | **Size (bp)** | **Distance (bp)** |
| C_162759 | DEL | 1 | 1047 | 178,080 |
| C_162750 | DEL | 1 | 387 | 196,131 |
| C_162646 | DEL | 1 | 911 | 828,066 |
| C_162621 | DEL | 1 | 91 | 997,792 |
| **Downstream of**<br>***DOT1L*** |  |  |  |  |
| C_162831 | DEL | 1 | 1282 | 183,339 |
| C_162851 | DEL | 1 | 1108 | 225,377 |
| C_162855 | DEL | 1 | 1029 | 268,689 |
| C_163022 | DEL | 1 | 905 | 918,230 |

**Table 2.**
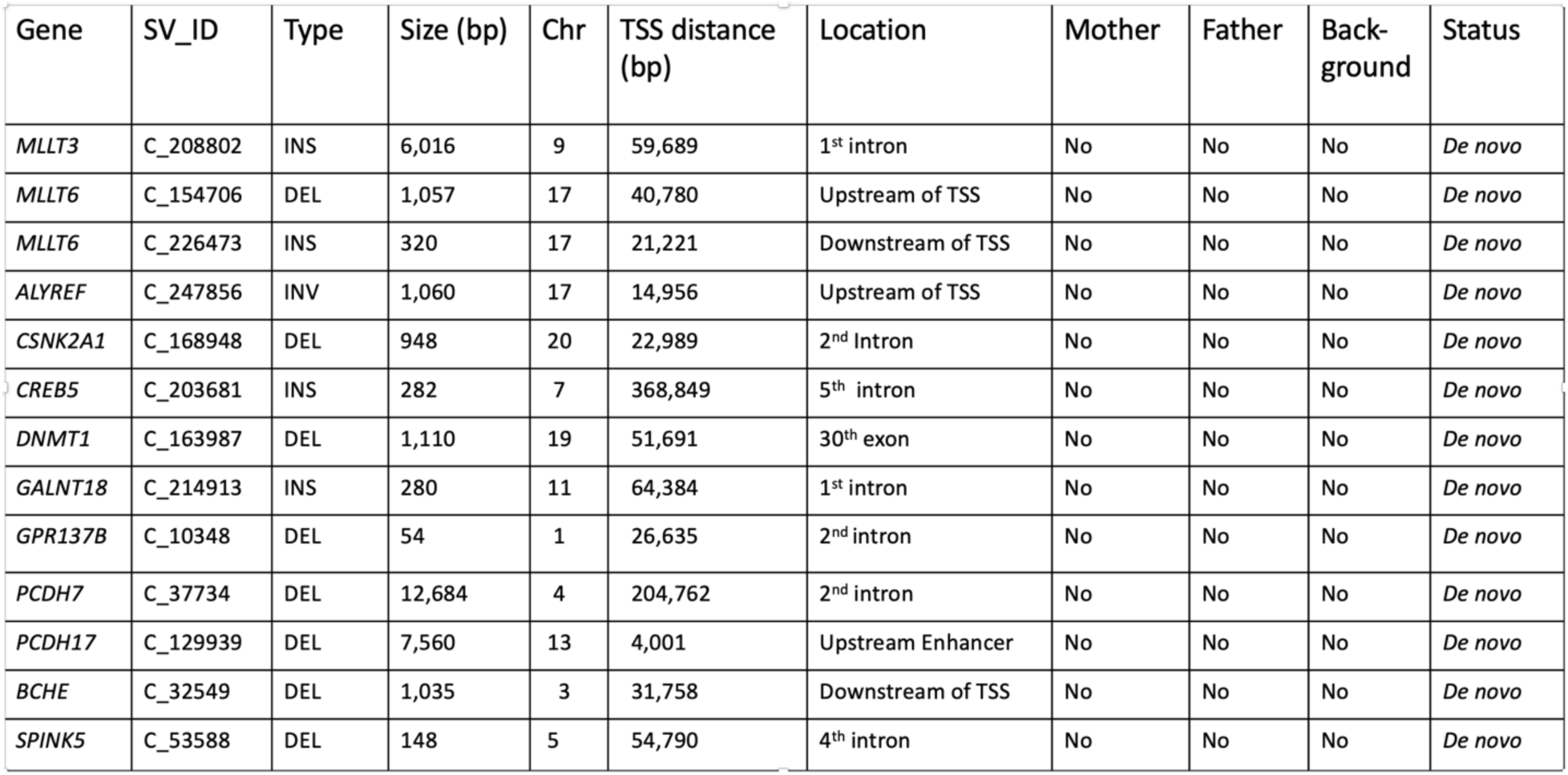
We identified five *de novo* SVs associated with four DOT1L subunits, *ALYFREF, CSNK2A1*, *MLLT3*, and *MLLT6*. Among DOT1L targets, we identified eight *de novo* SVs in *CREB5, DNMT1, GALNT18, GPR137B, PCDH7, PCDH17, BCHE*, and *SPINK5* in nsOFC probands.

| Gene | SV_ID | Type | Size (bp) | Chr | TSS distance (bp) | Location | Mother | Father | Back-ground | Status |
| --- | --- | --- | --- | --- | --- | --- | --- | --- | --- | --- |
| *MLLT3* | C_208802 | INS | 6,016 | 9 | 59,689 | 1<sup>st</sup> intron | No | No | No | *De novo* |
| *MLLT6* | C_154706 | DEL | 1,057 | 17 | 40,780 | Upstream of TSS | No | No | No | *De novo* |
| *MLLT6* | C_226473 | INS | 320 | 17 | 21,221 | Downstream of TSS | No | No | No | *De novo* |
| *ALYREF* | C_247856 | INV | 1,060 | 17 | 14,956 | Upstream of TSS | No | No | No | *De novo* |
| *CSNK2A1* | C_168948 | DEL | 948 | 20 | 22,989 | 2<sup>nd</sup> Intron | No | No | No | *De novo* |
| *CREB5* | C_203681 | INS | 282 | 7 | 368,849 | 5<sup>th</sup> intron | No | No | No | *De novo* |
| *DNMT1* | C_163987 | DEL | 1,110 | 19 | 51,691 | 30<sup>th</sup> exon | No | No | No | *De novo* |
| *GALNT18* | C_214913 | INS | 280 | 11 | 64,384 | 1<sup>st</sup> intron | No | No | No | *De novo* |
| *GPR137B* | C_10348 | DEL | 54 | 1 | 26,635 | 2<sup>nd</sup> intron | No | No | No | *De novo* |
| *PCDH7* | C_37734 | DEL | 12,684 | 4 | 204,762 | 2<sup>nd</sup> intron | No | No | No | *De novo* |
| *PCDH17* | C_129939 | DEL | 7,560 | 13 | 4,001 | Upstream Enhancer | No | No | No | *De novo* |
| *BCHE* | C_32549 | DEL | 1,035 | 3 | 31,758 | Downstream of TSS | No | No | No | *De novo* |
| *SPINK5* | C_53588 | DEL | 148 | 5 | 54,790 | 4<sup>th</sup> intron | No | No | No | *De novo* |

Previously, using *Dot1L* knockout mice, we analyzed the expression of DOT1L-dependent genes in mouse chondrocytes (Sutter et al., 2021). Among DOT1L target genes, we identified *de novo* SVs in *CREB5, DNMT1, GALNT18, GPR137B, PCDH7, PCDH17, BCHE,* and *SPINK5* that were present only in nsOFC probands, and absent in their parents (Table 2). We identified a 282 bp insertion (C_203681) in the fifth intron of *CREB5*. Similarly, we detected a 280 bp insertion (C_214913) in the first intron of *GALNT18*. We identified intronic deletions in *GRP137B* (C_10348, 54 bp), *PCDH7* (C_37734, 12,684 bp), and *SPINK5* (C_53588, 148 bp). Interestingly, a 1,110 bp deletion was detected in the 30^th^ exon of *DNMT1* (C_163987). We also identified a large 7,560 bp deletion (C_129939) in the promoter region of *PCDH17* and a 1,035 bp deletion (C_32549) in the downstream regulatory region of *BCHE*.

Surprisingly, we identified two children of African descent in the Kids First nsOFC datasets, KFG304 and KFG061, carrying a rare SV (C_248146) in *DOT1L* (Fig. 1). C_248146 represents a 6 kb inversion in the first intron of *DOT1L*. This inversion is rare because it occurs in only 34 of the 2981 background samples, corresponding to a minor allele frequency of approximately 1%. In the nsOFC Case–Parent Triads datasets, we detected six occurences among 722 samples, yielding an allele frequency of 0.83%.

**Figure 1.**
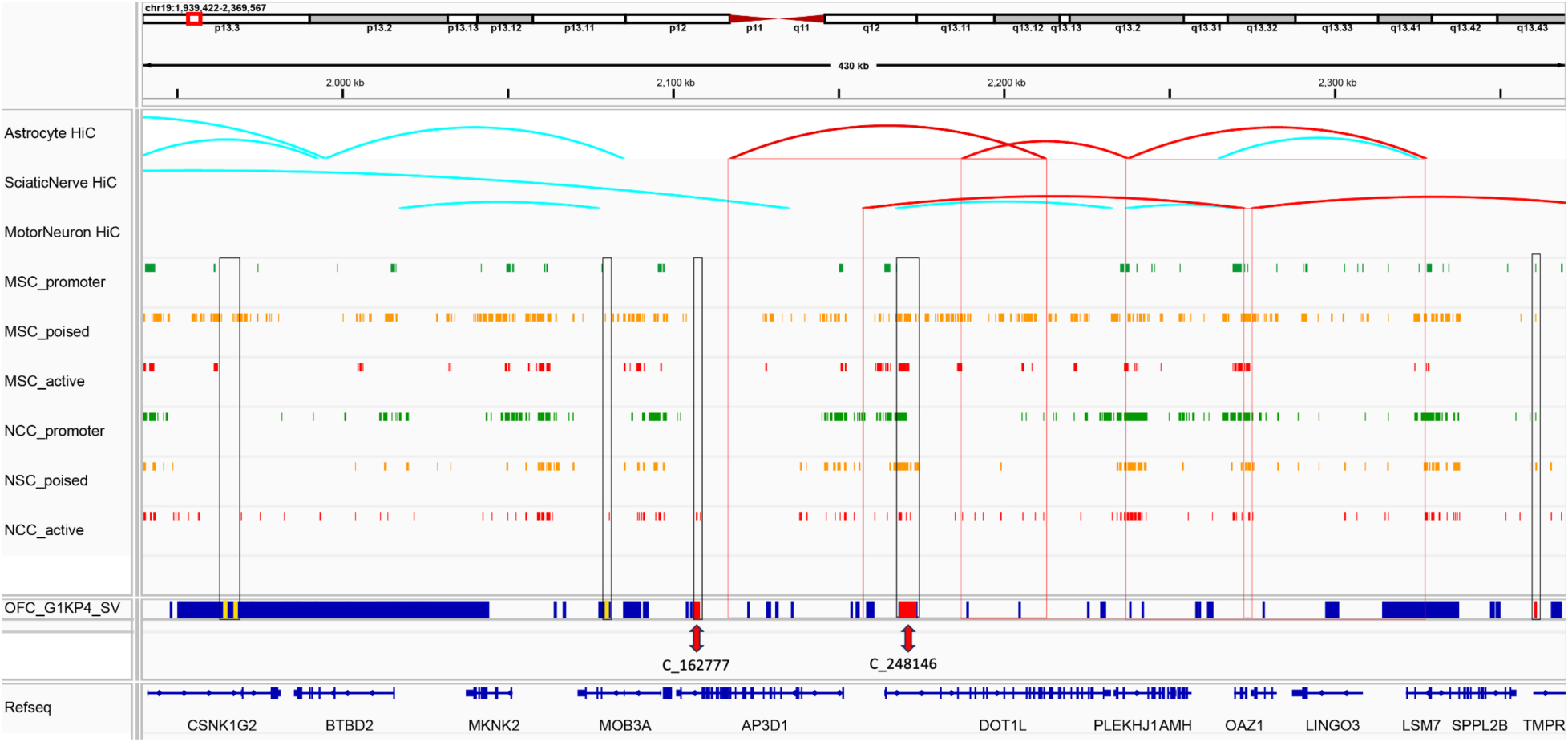
A rare SV of an active neural crest-specific enhancer candidate was identified as C_162777 (shown as a red bar). Craniofacial Atlas H3K27ac peaks for mesenchymal stem cells (MSC_active), show no overlap with ENCODE neural crest (NCC_active) H3K27ac peaks. This SV has an allele frequency of 0.0 in the 1000 Genomes population (Phase 4) and comes into contact with the *DOT1L* gene body, as shown by the Hi-C data in the top track (red line). A rare SV (C_248146 inversion, 6 kb) was identified in the first intron of *DOT1L* in two nsOFC children KFG304 and KFG061 (shown as a red bar). According to ENCODE data, this inversion is enriched in H3K27ac (NCC_active and MSC_active), H3K27me3, and H3K4me3 (NSC_poised and MSC_poised) histone marks. The region containing the C_248146 inversion is involved in Hi-C interactions with other genes (*PLEKHJ1*, *AMH*, and *OAZ1*).

In the KFG304 proband with CL/P, both parents exhibit a normal craniofacial phenotype and do not carry the inversion. In the KFG061 proband with cleft lip, both parents are phenotypically normal, although the mother carries the C_248146 inversion. According to ENCODE data, the C_248146 region is enriched for H3K27ac histone marks in NCCs and MSCs. In motor neurons, the region containing the C_248146 inversion participates in Hi-C interactions with *PLEKHJ1*, *AMH*, and *OAZ1*.

Additionally, we identified the Asian CL/P proband KF19056_01 proband carrying a rare SV (C_162777) upstream of *DOT1L* (Fig. 1). C_162777 represents 1 kb deletion encompassing a neural-crest-specific enhancer. Only two OFC samples (the parent and proband) carried this variant, and it had a frequency of less than 1% in the background population. Although both parents exhibit normal craniofacial features, the mother carries the same C_162777 deletion. The region within C_162777 exhibits properties of an active enhancer based on H3K27ac peaks in NCCs. Moreover, this region actively engaged in Hi-C interactions with the *DOT1L* locus in sciatic nerve cells drived from the neural crest lineage.

The conservation of these sequences in the healthy population, together with the presence of rare and *de novo* SVs in African and Asian nsOFC probands, indicates that the DOT1L complex and its targets comprise biologically important genes that warrant further investigation.

### Single-cell expression of *Dot1L* and its target genes in E12.5 mouse embryonic head mesenchyme

Based on differential expression of key developmental genes in snRNA-seq analysis, we defined 22 distinct cell clusters in the E12.5 mouse embryonic head (Fig. 2A). Mesenchymal and neural progenitor subtypes comprised the majority of embryonic head cells. We divided mesenchymal cells into nine clusters (clusters 1, 2, 3, 8, 12, 13, 14, 16, and 17) based on the expression pattern of the mesenchymal marker *Prrx1*. Differential expression of osteogenic genes (Fig. 2B) suggests that these mesenchymal progenitor populations represent osteogenic cells at various stages of differentiation. Consistent with this observation, critical genes associated with neural crest specification and differentiation, including *Twist2*, *Lef1*, *Satb2*, *Ednra*, *Tgfb3*, and *Pax3*, displayed differential expression patterns in the mesenchyme (Fig. 2B). Within cluster 5, we identified a subpopulation of dental epithelium cells enriched for *Lef1*, *Satb2*, *Tgfbr3*, and *Pax3*. *Dot1L* exhibited a wide range of expression in the embryonic head including mesenchyme (clusters 1, 2, 3, 8, 12, 13, 14 and 17) and dental epithelium (cluster 5).

**Figure 2.**
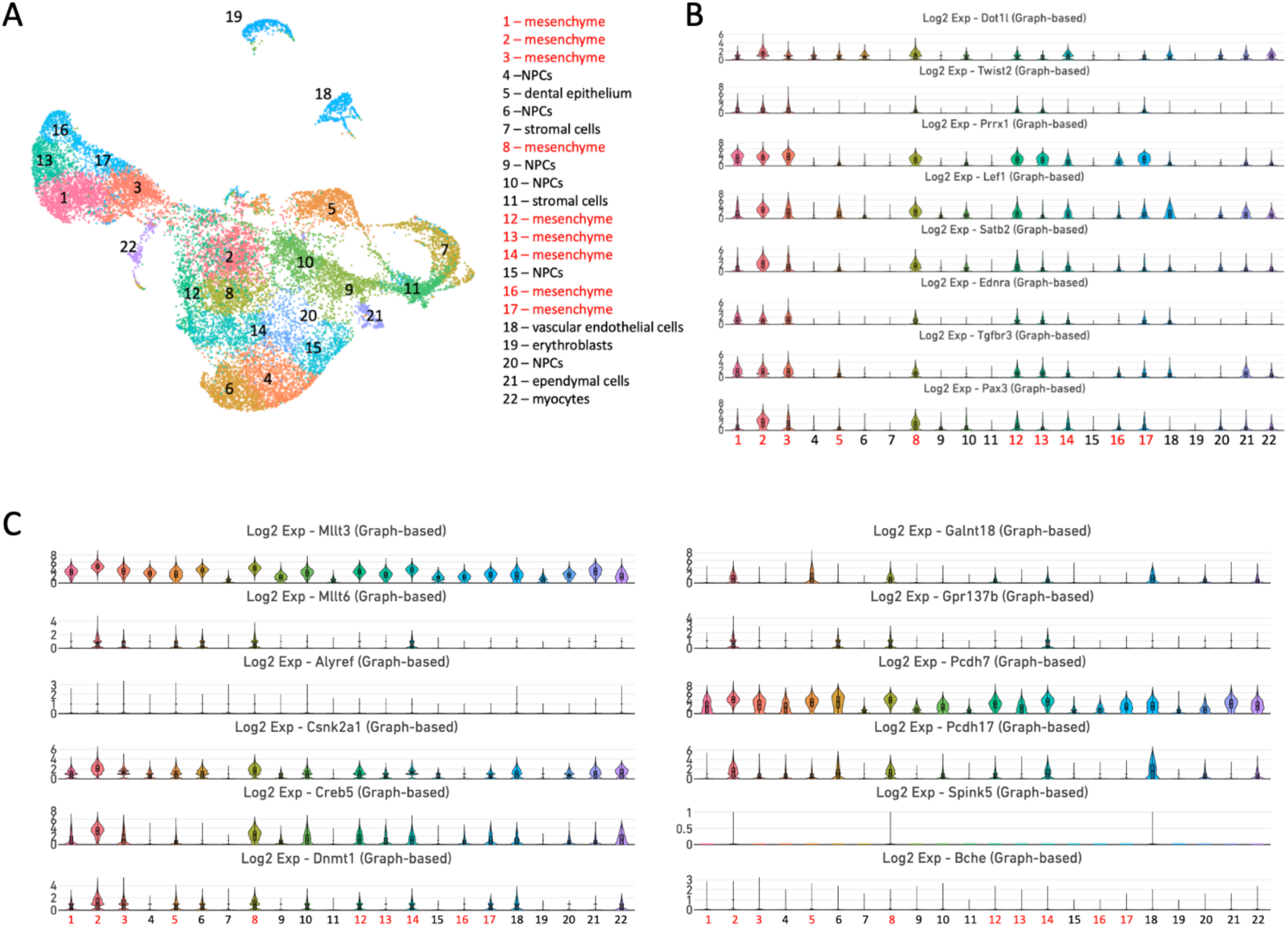
Differential expression of *Dot1L* and osteogenic genes in the E12.5 mouse embryonic head. Analysis of key developmental gene expression identified 22 distinct cell clusters based on snRNA-seq analysis. The majority of the embryonic head cell population consisted of several subtypes of mesenchyme and neural progenitor cells. We divided mesenchyme into nine clusters (clusters 1, 2, 3, 8, 12, 13, 14, 16, and 17) based on the expression pattern of the mesenchymal marker *Prrx1*.

We further analyzed snRNA-seq data to determine the expression of DOT1L subunit genes and targets identified in Table 2. *Mllt3*, *Csnk2a1*, and *Pcdh7* were broadly expressed throughout the embryonic head, including the *Dot1L*-positive mesenchyme (Fig. 2C). *Mllt6* was co-expressed with *Dot1L* in mesenchymal clusters 2, 3, 8, and 14. *Creb5* and *Dnmt1* were enriched in mesenchymal clusters 1, 2, 3, 8, 12, 14, and 17. *Pcdh17* was co-expressed with *Dot1L* in mesenchymal clusters 2, 3, 8, 12, and 14, whereas *Spink5* and *Bche* demonstrated modest expression.

### Co-expression of *Dot1L* with key orofacial genes in the mouse embryonic palate

We analyzed scRNA-seq data (Yan et al., 2024) from mesenchymal cells isolated from mouse palatal shelves at E12.5, E13.5, E14.0, and E14.5. Our analysis demonstrated that *Dot1L* is co-expressed with key transcription factors involved in palatogenesis (Figure 3A,B). Specifically, *Dot1L* was expressed in subcluster 1, which is enriched for genes critical for palate formation, including *Meox2, Twist1, Pou3f3, Dlx1, Sox9, Sox11, Foxf1, Foxp2, Lhx6, Lhx8, Trps1, Lef1, Tcf4, Shox2, Pax9,* and *Barx1*.

**Figure 3.**
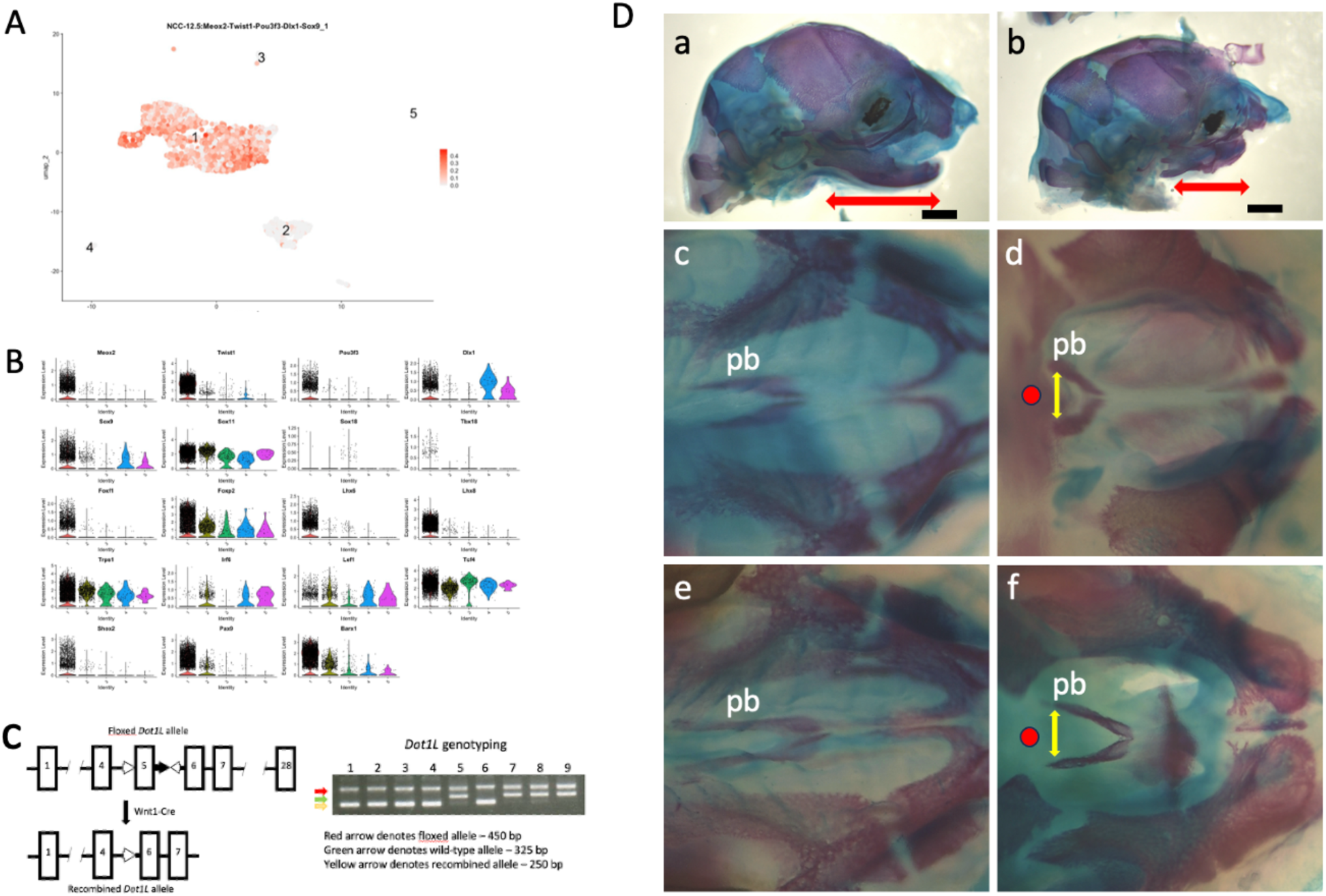
We identified five subclusters of palatal mesenchyme at E12.5 enriched in key osteogenic regulators. (A) *Dot1L* expression was primarily observed in subcluster 1, which includes genes essential for craniofacial development and palate formation. (B) *Dot1L* is co-expressed with key transcription factors critical for palate formation, including *Meox2, Twist1, Pou3f3, Dlx1, Sox9, Sox11, Foxf1, Foxp2, Lhx6, Lhx8, Trps1, Lef1, Tcf4, Shox2, Pax9,* and *Barx1*. (C) Generation of the conditional *Dot1L^fl/fl^* mouse line. (D) Wholemount images of E18.5 skull skeletal preparations stained with Alizarin red and Alcian blue. Wild-type embryos are shown in (a,c,e); *Dot1L^fl/fl^;Wnt1-Cre* mutant embryos are shown in (b,d,f). The double-arrowed red lines show the dramatic difference in mandibular lengths (b). Scale bar − 1mm. Reduced ossification of the palatine bones (pb) and absence of rugae bands were observed in *Dot1L^fl/fl^; Wnt1-Cre* mutant embryos (d,f). The red dot highlights clefting of the hard palate in mutants. The double-arrowed yellow lines indicate increased spacing between the palatine bones at the cranial base.

### Deletion of *Dot1L* in mouse embryos

We conditionally inactivated *Dot1L* in cNCCs using *Dot1L^fl/fl^* and *Wnt1-Cre* mice. The *Wnt1-Cre* line is extensively used in studies of neural crest development and its derivatives (Lewis et al., 2013). *Dot1L^fl/fl^* mice carry loxP sites flanking exon 5, which encodes most of the H3K79 methyltransferase domain (Fig. 3C). We crossed female *Dot1L^fl/fl^* mice with the *Wnt1*-*Cre* mice, and the resulting *Dot1L^fl/+^;Wnt1-Cre* male offspring with *Dot1L^fl/fl^* females to produce conditional knockout *Dot1L* cKO mice. PCR analysis confirmed efficient deletion of exon 5, which encodes the histone methyltransferase domain (Fig. 3C). Skeletal preparations of E18.5 embryos revealed midfacial underdevelopment and severe micrognathia in *Dot1L* cKO mice compared with littermate controls (Fig. 3D). *Dot1L* cKO embryos exhibited smaller snouts and mandibles, along with increased skull rounding, likely due to abnormal cranial base growth (Fig. 3D(b)). *Dot1L* cKO embryos also exhibited a cleft, accompanied by reduced ossification of the palatine bones and loss of rugae bands (Fig. 3D(d,f)), which were readily apparent in wild-type controls (Fig. 3D(c,e)).

### Rare SVs in *DOT1L* and its downstream target genes

We identified several families carrying SVs in *DOT1L* and downstream targets of DOT1L, including *SMOC2*, *ALDH1A*, and *CDH6*. Within two probands, KFG304 and KFG061, both carrying a 6 kb inversion in the first intron of *DOT1L* (C_248146), we also identified two SVs, C_66743 and C_66766, in *SMOC2* (Fig. 4A). C_66743 is a 1.7 kb deletion (KFG304), and C_66766 is a 1.4 kb deletion (KFG061), both located within intron 7 of *SMOC2*. In the KFG304 proband, neither the father (KFG305) nor the mother (KFG306) carries SVs in *DOT1L* or *SMOC2*, whereas the proband carries SVs in both genes. In the KFG061 family, both parents are phenotypically normal; however, the father (KFG062) carries the C_66766 deletion in *SMOC2* and the mother (KFG063) carries the C_248146 inversion in *DOT1L*. In the KF19056 family, the child (KF19056_01) carries SVs in *DOT1L*, *ALDH1A*, and *CDH6*, whereas the mother (KF19056_02) carries SVs in *DOT1L* and *ALDH1A1*, and the father (KF19056_03) carries only C_196287 in *CDH6* (Fig. 4A). snRNA-seq analysis of the E12.5 embryonic head showed that *Cdh6* is enriched in *Dot1L*-positive mesenchyme, including clusters 1, 2, 3, 8, 12, 13, 14, and 17, as well as dental epithelium (cluster 5). *Smoc2* is expressed in clusters 1, 2, 3, 8, and 12, whereas, *Aldh1a1* shows modest expression in clusters 2 and 8. During palatogenesis (Yan et al., 2024), *Smoc2, Aldh1a, Cdh6,* and *Dot1L* are co-expressed in subclusters 1 and 2 of palatal mesenchyme at E12.5 and E13.5 (Fig. 4C).

**Figure 4.**
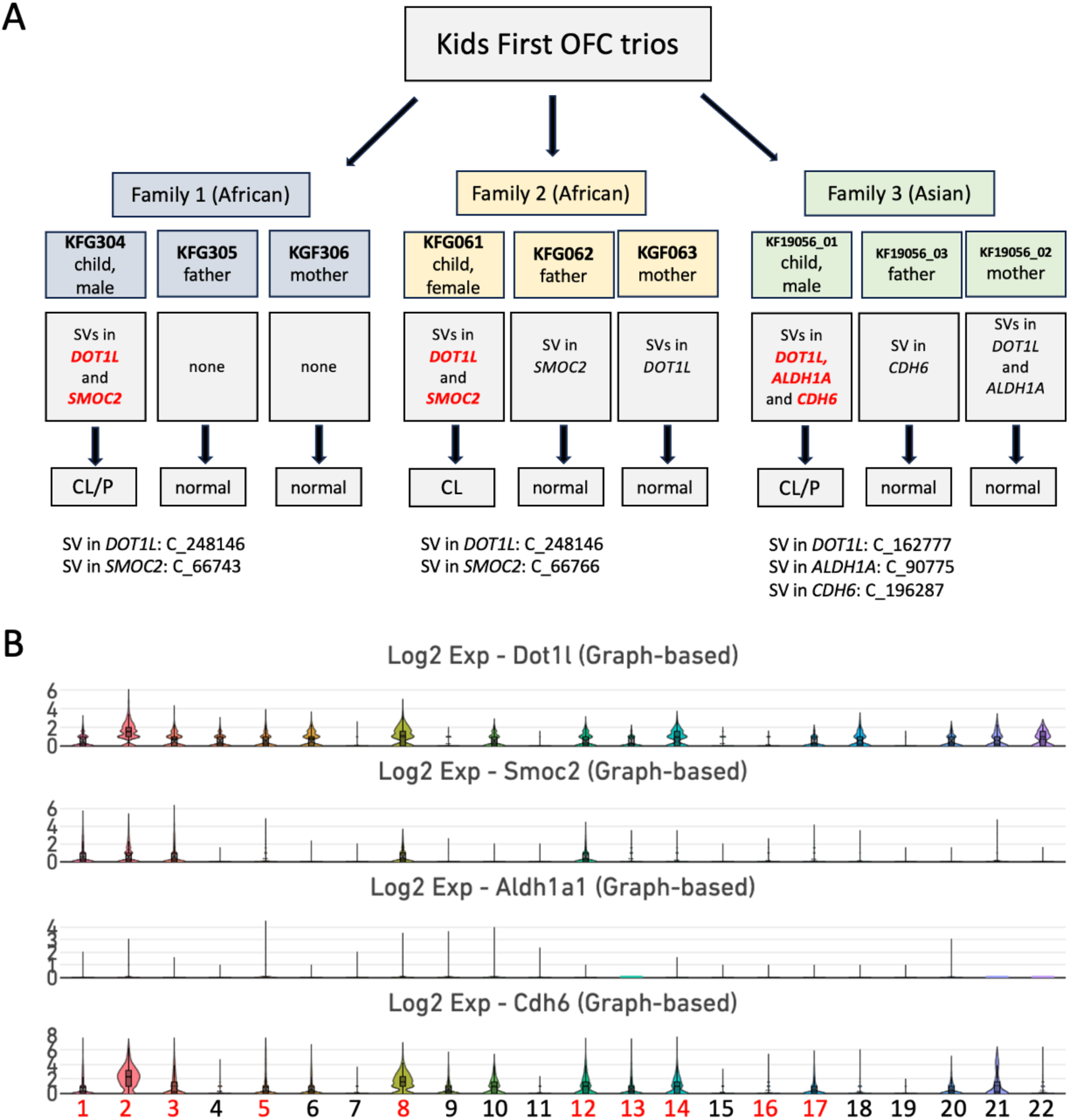
We identified two children of African descent in the Kids First nsOFC datasets, KFG304 and KFG061, carrying a rare SV (C_248146 inversion) in the first intron of *DOT1L*. We also identified rare SVs in *SMOC2* in the same two probands. In the KF19056 family, the child inherited SVs in *DOT1L* (C_162777) and *ALDH1A* (C_90775) from the mother (KF19056_02), and an SV (C_196287) in *CDH6* from the father (KF19056_03). (B) snRNA-seq of the E12.5 mouse heads showed that *Dot1L*, *Smoc2*, and *Cdh6* are co-expressed in mesenchymal cells (clusters 1, 2, 3, 8, and 12) which are enriched in osteogenic markers. *Aldh1a1* exhibits modest expression in the embryonic head.

## Discussion

Many factors including genetic predispositions and environmental interactions contribute to craniofacial abnormalities (Im et al., 2025; Shull and Artinger, 2024). Epigenetic mechanisms associated with histone modifications, DNA methylation, microRNAs, and noncoding RNAs play crucial roles during palatogenesis, as disturbances in chromatin structure can lead to a wide range of craniofacial defects, including cleft palate (Kano et al., 2022; Shpargel et al., 2020; Loenarz et al., 2010; Ulschmid et al., 2024; Shi et al., 2024; Iwaya et al., 2023).

The conservation of genes encoding DOT1L complex subunits in the healthy population, together with the presence of rare and *de novo* SVs in African and Asian nsOFC probands, indicates that the DOT1L complex comprises functionally important genes involved in craniofacial development. Furthermore, we identified several families carrying SVs in *DOT1L* and its target genes, suggesting that their combined effects may contribute to orofacial defects, including CL/P.

Previously, we showed that *Smoc2*, *Aldh1a1*, and *Cdh6* are downstream targets of DOT1L in mouse chondrocytes (Sutter et al., 2020). In two nsOFC children carrying a rare SV (C_248146) in the first intron of *DOT1L*, we also identified rare SVs (C_66743 and C_66766) in *SMOC2* (Fig. 4A). SMOC2 (Secreted Modular Calcium-Binding Protein 2) is a glycoprotein involved in calcium ion binding, extracellular matrix organization, and BMP2 signaling (Bloch-Zupan et al., 2011). Mutations in *SMOC2* are associated with dentin dysplasia type I (Bloch-Zupan et al., 2011). In mice, loss of *Smoc2* impairs bone healing and produces age-dependent bone loss (Morkmued et al., 2020). Interestingly, brachycephalic dogs with reduced expression of *Smoc2* exhibit facial retrusion, proximodistal shortening of the snout, and widening of the hard palate (Marchant et al., 2020).

In the KF19056 family, the child inherited SVs in *DOT1L* (C_162777) and *ALDH1A* (C_90775) from the mother (KF19056_02), and an SV (C_196287) in *CDH6* from the father (KF19056_03) (Fig. 4A). *Cdh6* is expressed in NCCs migrating from the neural tube in mouse embryos (Inoue et al., 1997). During epithelial−mesenchymal transformation, *Cdh6* promotes NCC detachment through regulation of F-actin dynamics (Clay and Halloran, 2014). Mutations in *CDH1*, a close paralog of *CDH6*, are associated with an increased risk of CL/P (Frebourg et al., 2006; Brito et al., 2016).

*ALDH1A1*, together with *ALDH1A2* and *ALDH1A3*, belongs to the retinaldehyde dehydrogenase family involved in retinoic acid signaling (Duester, 2022). Excess of retinoic acid leads to cleft palate in both human and mice (Okano et al., 2014). In hepatocellular carcinoma, ALDH1A1 stabilizes GLI2 and enhances Hedgehog signaling activity (Yan et al., 2016). Individuals carrying truncating mutations in GLI2 displayed midface hypoplasia, CL/P, and hypothelorism (Bear et al., 2014). Likewise, *Gli2* mutant mice develop cleft palate, tooth defects, and shortened limbs (Mo et al., 1997). Notably, depletion of LRP6, a co-receptor of WNT/β-catenin signaling, leads to CL/P associated with ectopic expression of *Aldh1a3* (Song et al., 2009).

Our findings suggest that the cumulative effects of SVs in *DOT1L* and its downstream target genes contribute to the nsOFC phenotype, supporting an oligogenic model in which the concurrent presence of multiple SVs promotes craniofacial malformations. This interpretation is consistent with emerging oligogenic frameworks proposed in recent studies (Almansoori et al., 2025). We hypothesize that *DOT1L* and its downstream target genes act within the same gene regulatory network during palatogenesis. Understanding the crosstalk between H3K79 methylation and DOT1L targets is important for identifying disease−susceptibility genes, as the combined effects of SVs may be more pronounced than the effects of individual variants alone. These findings provide a testable framework that can guide future mechanistic studies and experimental validation.

In conclusion, our study advances understanding of large genomic alterations and their potential contributions to orofacial cleft risk. We established the importance of DOT1L in craniofacial development and identified *Smoc2*, *Aldh1a1*, and *Cdh6* as candidate contributors to the etiology of nsOFCs. Additionally, other DOT1L target genes may represent promising candidates for future investigations.

## Author contributions

T.B. and D.S. initiated the study conception and design. T.B. generated and analyzed the computational data. D.B designed experiments. Material preparations, data collection, and data analyses were performed by T.B., D.B., and D.S.. D.B. wrote the main manuscript text. All authors critically revised and approved the final version of the manuscript.

## Data availability

The data that support the findings of this study are available from the corresponding author upon request.

